# Statistical analysis plan for the Identification and Treatment of Hypoxemic Respiratory Failure (HRF) and ARDS with Protection, Paralysis, and Proning: a type-1 hybrid stepped-wedge cluster randomized effectiveness-implementation study

**DOI:** 10.1101/2023.03.17.23287218

**Authors:** Ken Kuljit S. Parhar, Andrea Soo, Gwen Knight, Kirsten Fiest, Daniel J. Niven, Gordon Rubenfeld, Damon Scales, Henry T. Stelfox, Danny J. Zuege, Sean Bagshaw

## Abstract

**Background:** The Identification and Treatment of Hypoxemic Respiratory Failure (HRF) and ARDS with Protection, Paralysis, and Proning (TheraPPP) study is a type-1 hybrid stepped-wedge cluster randomized effectiveness-implementation study involving 17 adult Intensive Care Units (ICUs). This study will evaluate the effectiveness and implementation of an evidence-based, stakeholder-informed, multidisciplinary care pathway called *Venting Wisely* that standardizes the diagnosis and delivery of life-saving therapies for critically ill patients with Hypoxemic Respiratory Failure (HRF) and acute respiratory distress syndrome (ARDS).

**Objective:** To describe a pre-specified statistical analysis plan (SAP) for the TheraPPP study prior to completion of recruitment, electronic data retrieval, and before any analysis has been conducted.

**Methods and analysis:** The Statistical Analysis Plan (SAP) was designed by the principal investigators and senior biostatistician and reviewed in detail by the *Venting Wisely* Scientific Steering Group before being approved. This statistical analysis plan is reported in accordance with Guidelines for the Content of Statistical Analysis Plans in Clinical Trials. A study specific CONSORT diagram and baseline characteristics table were developed. We estimate a total of 18816 mechanically ventilated patients will be included in this study with 11424 patients pre-implementation and 7392 patients post implementation. Given that ARDS patients are an important subgroup within this study, we estimate that this will generate a sample size of 2688 sustained ARDS patients within our TheraPPP study cohort. The primary clinical outcome is 28-day ventilator free days (VFDs). For the primary analysis, we will compare the mean 28-day VFDs pre-implementation and post-implementation using a mixed effects linear regression model to account for clustering of patients within site. Secondary clinical outcomes will be similarly compared pre-implementation and post-implementation using mixed effects linear or logistic regression models, as appropriate. All models will be adjusted for age, sex, severity of illness (sequential organ failure assessment score on admission) and severity of hypoxemia on admission based on PF ratio, as well as type and size of ICU. Pre-specified subgroups will include patient sex, age, HRF, ARDS, Covid-19 and cardiac surgical status, body mass index (BMI), height, illness acuity, and ICU volume.

**Ethics and Trial Registration:** The study has received ethics approval from the University of Calgary (20-0646) and the University of Alberta (pro00112232). The trial was registered with ClinicalTrials.gov (NCT04744298) prior to the enrollment of any patients on Feb 8, 2021.

## 3 Administrative Information

This document has been written based on information contained in the study protocol version 2.5, dated February 14, 2023 in accordance with Guidelines for the Content of Statistical Analysis Plans in Clinical Trials.(1) Study methods will be conducted and reported in accordance with standards for reporting stepped wedge cluster randomised trials (CONSORT, SW-CRT extension),(2) and standards for reporting implementation studies(StaRI)(3) and their replication (TIDieR).(4) Qualitative work will be reported using Standards of Reporting of Quality Research guidelines (SRQR) and Consolidated criteria for Reporting Qualitative research (COREQ).(5, 6) The protocol is also reported in accordance with the Standard Protocol Items: Recommendations for Interventional Trials (SPIRIT) guidance and checklist 2013.(7)

The study has received ethics approval from the University of Calgary (20-0646) and the University of Alberta (pro00112232). The study protocol is registered on clinicaltrials.gov NCT04744298.

## 4 Introduction

### 4.1 Background and rationale

Hypoxemic respiratory failure (HRF) and acute respiratory distress syndrome (ARDS) are common conditions among patients admitted to the intensive care unit (ICU). Treatment of the patients is complex. Evidence-based therapies that improve survival exist; however, implementation is inconsistent and variable. The Institute of Medicine has recommended standardized care processes to improve the reliability and safety of care.(8) We developed the *Venting Wisely* pathway to reduce practice variation and improve adherence to evidence-informed therapy. A study is needed to evaluate effectiveness, cost effectiveness, and implementation of the pathway.

### 4.2 Study objectives and hypothesis

The overall objective of this study is to improve the quality of care for patients with HRF by implementing a rigorously developed, evidence-based, stakeholder-informed, multidisciplinary standardized care pathway called *Venting Wisely* that standardizes the diagnosis and delivery of life-saving therapies for critically ill patients with HRF.

The specific objectives are to evaluate:

1. **Clinical Effectiveness** of the pathway using a pragmatic registry-based cluster randomized stepped-wedge implementation study involving 17 adult ICUs.
2. **Implementation** of the pathway by conducting a process evaluation which will assess the **fidelity** of the delivered interventions and clinician perceptions about the **acceptability** of the pathway.
3. A **cost-effectiveness** analysis of the pathway.

We *hypothesize* that the pathway will increase adherence to life-saving therapies, improve patient outcomes, and save costs within the health care system.

## 5 Study Methods

### 5.1 Study design

The study is designed as an effectiveness-implementation hybrid study design (type 1).(9) This study design evaluates both clinical e*ffectiveness* and *implementation* of the pathway, but is primarily powered to the primary clinical effectiveness outcome. Implementation will occur via a pragmatic registry-based stepped wedge cluster randomized implementation study.(9)

### 5.2 Setting

The study will be conducted at 17 adult ICUs in Alberta, Canada. These 17 ICUs comprise a mix of tertiary, community, and rural ICUs. One ICU (Calgary) served as the setting for a pilot study (completed September 2020). The remaining 16 ICUs will participate in the full study.

### 5.3 Randomization

The unit of randomization will be a cluster. Two ICUs will comprise each cluster. Each ICU will be randomly assigned to one of the 8 clusters to initiate the intervention at different times according to the stepped wedge allocation schedule (See Figure 1). Sites will be randomized using computer generated random number sequence by a blinded investigator. Details of the randomization method are held securely in the statistics master file. Two sites will be selected at any time. ICU sites will be deferred from a randomization step if critical unreadiness events are identified which would include Covid-19 related capacity strain, transition to a new electronic health record, or undergoing Provincial ICU accreditation. Sites will be randomized and notified four to eight weeks prior to the initiation schedule to prevent contamination.

**Figure 1.**
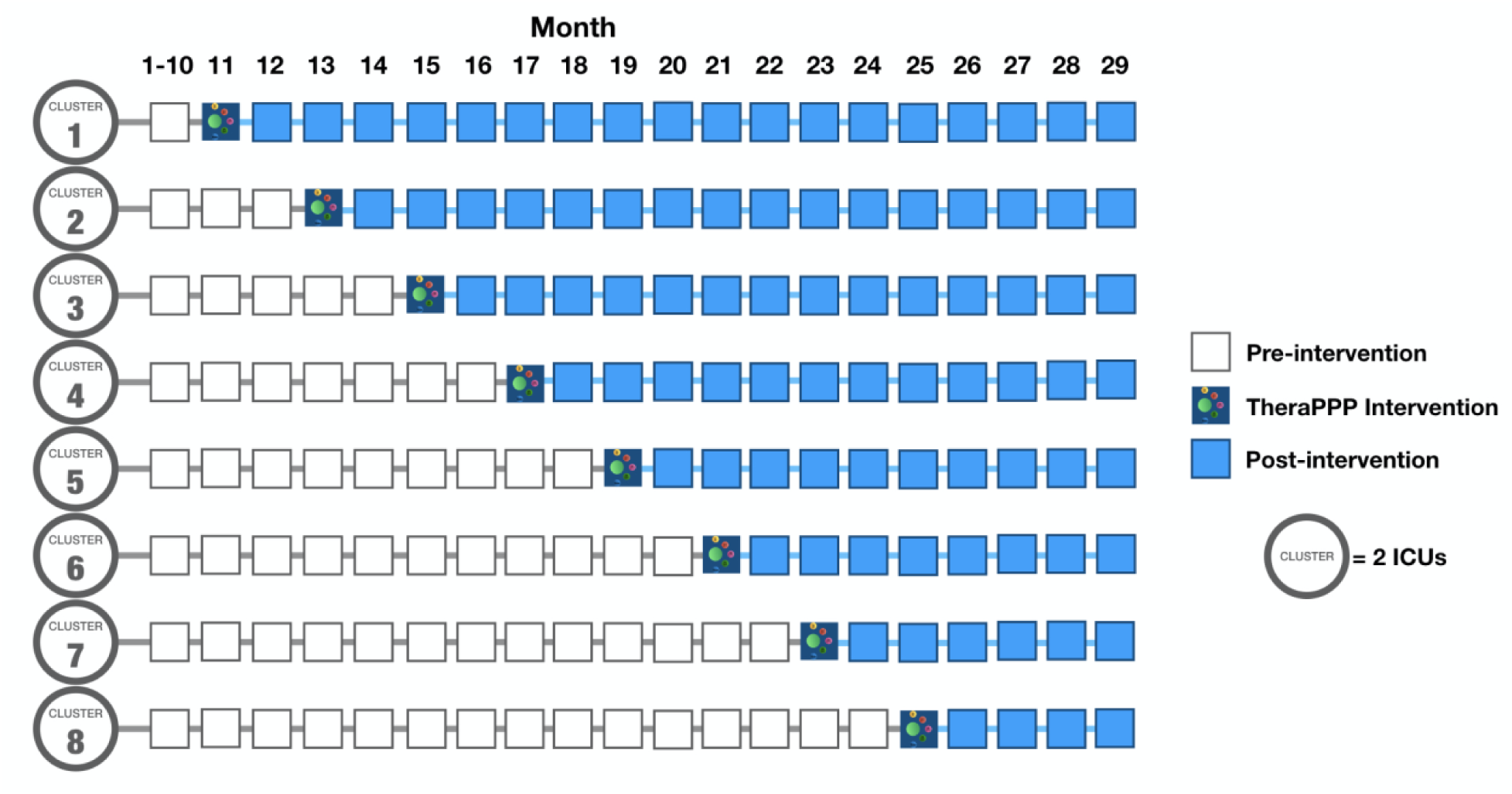
TheraPPP Study: stepped wedge cluster randomization

### 5.4 Study Duration

There will be a 10-month (June 2020 - March 2021) baseline data collection period at the beginning of the study common to all sites. Baseline data will continue to be collected until the intervention is implemented in an ICU. The intervention will be implemented into one cluster (two ICUs) every two months. The first month of each step will be a transition period from usual care, during which data will not be analyzed. Once implemented, the cluster will continue to receive the intervention for the remainder of the study. There will be a four month follow up period after implementation of the final cluster. The total study duration will be 29 months (June 2020-October 2022).

### 5.5 Sample Size

This study will assess both the effectiveness and implementation of a standardized management pathway for HRF and ARDS. The study is a type 1 hybrid effectiveness-implementation design and therefore is primarily powered for the effectiveness outcome.(9) We also provide sample size calculations for the implementation outcomes to estimate the effect sizes and precision of estimates that can be detected.

#### 5.5.1 Clinical Effectiveness Sample Size

The design of the cluster randomized stepped wedge TheraPPP trial incorporated several considerations. The study balances detection of a meaningful clinical difference in the primary outcome of 28-day ventilator free days (VFDs), with a pragmatic and efficient implementation of the pathway. A step duration that was too long would potentially result in contamination or secular changes in practice. A step duration that was too short would not allow adequate time for implementation of the pathway within each cluster. The number of ICUs per cluster also balanced the study team’s ability to implement the pathway in a given step. Too many ICUs per cluster would not be feasible for the implementation team, but alternatively, too few would result in a study duration that was too long and also susceptible to contamination or secular changes in practice.

Based on the considerations above, the final study design included a ten-month baseline data collection period, eight clusters with two ICUs per cluster, and implementation of the pathway in one cluster every two months followed by a four month post implementation period following the last cluster. Based on historical ICU admission rates in Alberta from 2018-2019 (unpublished eCritical registry data), we estimate a total of 18816 mechanically ventilated patients will be included in this study with 11424 patients pre-implementation and 7392 patients post implementation. Based on this, a baseline mean VFDs of 21 (standard deviation (SD) 10, intraclass correlation coefficient (ICC) = 0.15), a 90% power and a two-sided α=0.05 we estimate an ability to detect a difference of 0.9 VFDs (see Table 1).

**Table 1.**
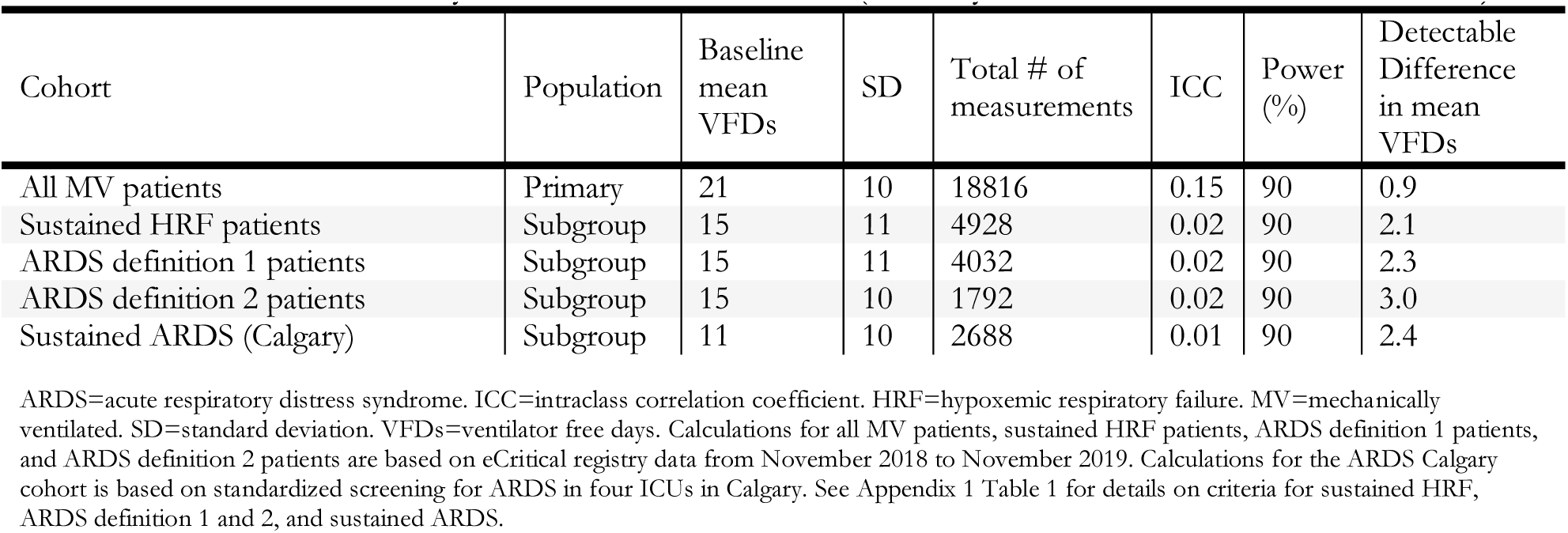
Ventilator Free Days Detectable Differences (Primary Clinical Effectiveness Outcome)

Given that ARDS is an important subgroup of patients within this cohort that would receive most steps of the pathway, we also wanted to ensure that we would recruit enough patients from this subgroup of interest over the study duration. To estimate the ARDS population within this cohort, we applied a population-based incidence of ARDS that was derived within Calgary using standardized screening for ARDS.(10) Using this historical population-based incidence, we anticipate an average of 12 sustained ARDS patients (see Appendix Table 1 for definition) per 2-month period per site (based on our observed sustained ARDS incidence of 0.42 per bed per month in Calgary).(10) Based on the stepped wedge design, we estimate that this will generate a sample size of 2688 sustained ARDS patients within our TheraPPP study cohort. This number of patients will provide the ability to detect a minimum difference of 2.4 days (11 to 13.4) in the mean 28-day VFDs (with a 90% power and a two-sided α=0.05, ICC = 0.01) within this subgroup. The minimal clinically important difference of 2.4 days is similar to other ARDS trials.(11-13) The ICC was estimated to be 0.011 (95% confidence interval (CI) 0.00-0.20) based on our previous epidemiological description of patients with sustained ARDS which was based on four sites in Alberta.(10) The VFD effect difference in ARDS patients that this study is powered to is conservative, and targets the lower limit of the pooled effect difference observed in our previously published systematic review on the use of standardized management pathways for HRF and ARDS (standardized mean difference increase of 3.48 (2.43-4.54) days).(14)

To improve the reliability of these estimations we conducted several sensitivity analyses. In order to ensure these assumptions were applicable to the cohort of 16 ICUs, particularly the ICCs that we are using to estimate our detectable difference, we conducted an alternative estimation based on provincial eCritical registry data from 2018-2019. Given that all mechanically ventilated patients are eligible for the pathway but not all patients may receive all elements, we also estimated the sample size based on recent data from 2018-2019 using a more liberal approach to eligible patients that would be estimated based upon patients with sustained HRF (see Appendix Table 1 and Table 1). The detectable difference was similar to our previous estimates. We also examined the proportion of patients with ARDS, as well as the detectable difference in VFDs using two registry-based definitions of ARDS (ARDS definition 1 and definition 2, see Appendix Table 1 & 4). ARDS is not formally or routinely documented in day-to-day electronic health records and therefore a registry based method was used. The diagnosis of ARDS by this method may be less precise; however, was used to provide an ICC for all 16 ICUs. Using these alternative assumptions, the minimum detectable difference would be similar (see Table 1). The power calculation was performed using the Stata function “steppedwedge”.(15, 16)

#### 5.5.2 Implementation Sample Size

Implementation will be assessed by the fidelity to the intervention and by the acceptability of the pathway to the healthcare team.

##### Fidelity

Given this is a type 1 hybrid study, we also estimated the detectable difference in our primary implementation outcome (Composite Fidelity Score [CFS%]). Based on our primary implementation outcome of CFS%, and using a baseline CFS of 20%, a standard deviation of 32%, 18816 patients, ICC of 0.31, (with a 90% power and a two-sided α=0.05), we estimate the study could detect a difference of 2.6% in mechanically ventilated patients (See table 2). We conducted similar sensitivity analyses for the sample size using recent data from 2018-2019 from the eCritical registry using a more liberal approach to eligible patients that would include based upon patients with sustained HRF and registry-based definitions of ARDS (Table 2). Estimates for sustained HRF or using registry-based ARDS definitions were similar (Table 2). Based on the sample of 2688 sustained ARDS patients (calculated for the primary clinical outcome) and a baseline mean CFS% of 56% (SD of 29%), this study would have power to detect a minimum difference of 7.1% (56% to 63.1%) in the mean CFS score (with 90% power and a two-sided α=0.05, ICC=0.02). This difference was believed to represent a clinically important difference, as a similar improvement was observed in our pilot intervention. Despite the pilot study not being powered for clinical outcomes, this degree of improvement was associated a reduction in driving pressure, mechanical power, and ICU length of stay (LOS). The power calculation was performed using the Stata function “steppedwedge”.(15, 16)

**Table 2.** Composite Fidelity Score Detectable Differences (Primary Implementation Outcome)

| Cohort | Population | Baseline mean CFS (%) | SD (%) | Total # of measurements | ICC | Power (%) | Detectable Difference in mean CFS (%) |
| --- | --- | --- | --- | --- | --- | --- | --- |
| All MV patients | Primary | 20 | 32 | 18816 | 0.31 | 90 | 2.6 |
| Sustained HRF patients | Subgroup | 35 | 29 | 4928 | 0.32 | 90 | 4.6 |
| ARDS definition 1 patients | Subgroup | 36 | 30 | 4032 | 0.33 | 90 | 5.2 |
| ARDS definition 2 patients | Subgroup | 38 | 30 | 1792 | 0.38 | 90 | 7.4 |
| Sustained ARDS (Calgary) | Subgroup | 56 | 29 | 2688 | 0.02 | 90 | 7.1 |

##### Acceptability (Surveys)

We estimate up to a total of 1000 survey responses from clinicians. Based on our pilot study and previous work (5) we anticipate a conservative response rate of 50% (625 surveys completed of 1250 distributed) which will provide 95% binomial confidence intervals of ±3.9%.

### 5.6 Framework

This study is designed as a superiority hypothesis testing framework.

### 5.7 Interim analysis

Interim analyses are not planned and will not be performed.

### 5.8 Timing of final analysis

The primary analysis will be prepared once all patients have reached 90-days of follow-up (with time=0 being the initiation of mechanical ventilation). Final electronic data will be available within six months of the 90-day follow-up period (see Figure 2). Completion of final analysis is targeted for October 2023). This statistical analysis plan version 1 (February 22, 2022) was added to clinicaltrials.gov and posted publicly on a preprint server (medrxiv.org) prior to the retrieval of electronic data and before any analyses had been conducted.

**Figure 2.**
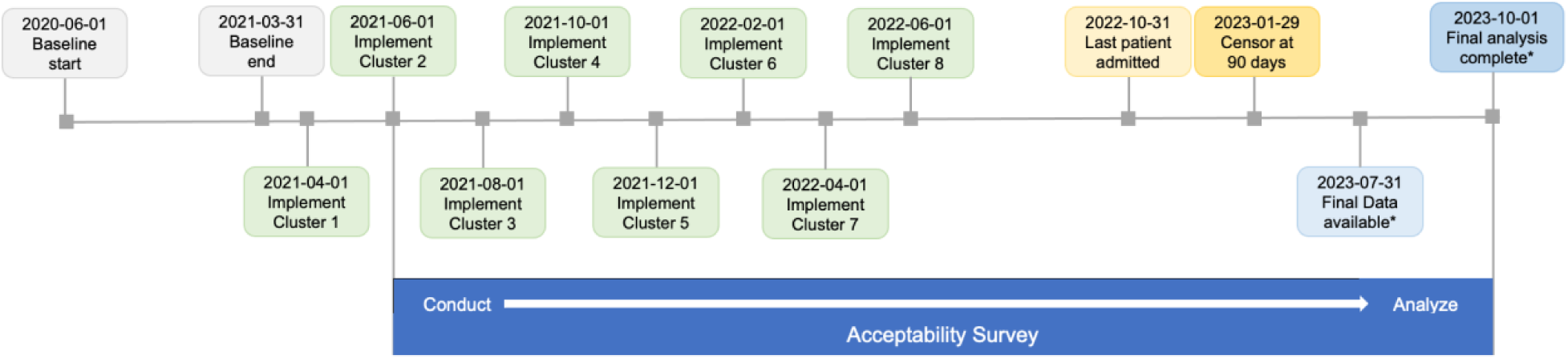
Study Timeline *=Estimated date

### 5.9 Timing of outcome assessments

Timing of outcome assessments for primary outcomes are listed below and all outcomes (primary, secondary, and exploratory) are detailed in section 8.1 and Appendix Table 3.

#### 5.9.1 Clinical effectiveness

For the primary outcome of 28-days VFDs, which is a composite outcome of survival and days spent not ventilated over the first 28 days, *timing* is measured as follows:

- The first day of ventilation is day 0
- The end date is the date of first ventilation + 28 days (censored at hospital discharge)

VFDs are calculated as previously described (17, 18):

- 0 if the patient dies within 28 days of mechanical ventilation
- 0 if the patient is still being ventilated after 28-days following initiation of mechanical ventilation whether they are extubated and survive or die after this timepoint
- 28 – x (if the patient successfully liberated from ventilation x days after initiation, within the 28-day timeframe)
- If a patient is invasively mechanically ventilated via endotracheal tube or tracheostomy for any period of time in a 24-hour period (0000-2359) this is considered a ventilated day
- In the case of repeat intubation episodes, liberation will be counted from the day of final successful extubation

#### 5.9.2 Implementation

##### Fidelity

The primary outcome of adherence is the CFS. The individual metrics of the CFS are measured per admission (height) and daily. See *Implementation Outcomes – Fidelity Indicators* section in Appendix Table 3 for details.

##### Acceptability (survey)

To evaluate acceptability outcomes, invitations to participate in the *acceptability survey* will be sent to clinicians (nurses, physicians, and Respiratory Therapists) two to six months post implementation in each cluster.

## 6 Statistical Principles

### 6.1 Confidence intervals and P values

The threshold for the entire analysis of primary and secondary outcomes will be two-sided using a 5% significance level (α=0.05). Measures of association will be reported using difference in means or odds ratios with 95% CI as appropriate. There is only one primary clinical effectiveness outcome, therefore no adjustment for multiplicity is required. For secondary outcomes we will the report the false discovery rate to account for multiplicity of testing.

### 6.2 Adherence and protocol deviations

#### 6.2.1 Adherence

Fidelity of the intervention will be tracked using five evidence-based process of care indicators that reflect the five key steps of the pathway which are routinely charted in the electronic health record:

1. Proportion of patients ventilated with a height measured (step 1)
2. Proportion of eligible patient days who receive a tidal volume ≤8mL/kg predicted body weight (step2/3)
3. Proportion eligible patient days who have a plateau pressure measured (step 3)
4. Proportion of eligible patient days who receive neuromuscular blockade (step 4)
5. Proportion of eligible patient days who receive prone ventilation (step 5)

The CFS awards points for the five indicators above that are met and provides an overarching indicator of adherence. See 8.1.2 and the *Implementation Outcomes – Fidelity Indicators* section in Appendix Table 3 for additional details of adherence indicators.

Adherence will be presented pre and post implementation of the intervention (Mean, Median, interquartile range (IQR), p-value). Time trends in the CFS will also be presented for all mechanically ventilated patients, patients with HRF, and patients with ARDS (definition 2, see Appendix Table 1 for definition). Fidelity process of care indicators will also be used to improve pathway adherence through monthly audit and feedback reports.

#### 6.2.2 Protocol deviations

The following protocol deviations will be summarized:

1. An ICU site withdraws from implementation of the pathway.
2. An ICU is not able to initiate implementation on their scheduled start date.
3. An ICU is not able to chart the requisite data required for audit and feedback, implementation outcomes and clinical outcomes (e.g., due to pandemic surge crisis charting).

### 6.3 Analysis populations

We will analyse the data using an intention to treat analysis. In the event of a patient moving from an intervention site to a non-intervention site, see section 7.4 for details.

## 7 Trial Population

### 7.1 Screening Data

All patients admitted to the adult ICU will be screened daily for eligibility for the pathway.

### 7.2 Eligibility

All mechanically ventilated patients admitted to the ICU will be included in the study and receive the pathway intervention. There are no exclusion criteria for entry into in the pathway; however, not all steps will be applicable to all mechanically ventilated patients. Patients cared for in non-traditional ICU settings due to expanded Covid-19 surge capacity (e.g. Coronary Care Unit, post-operative care units) will be included.

### 7.3 Recruitment

Number of ICUs, number of eligible patients, and exclusions will be detailed in the CONSORT flow diagram. See Figure 3.

**Figure 3.**
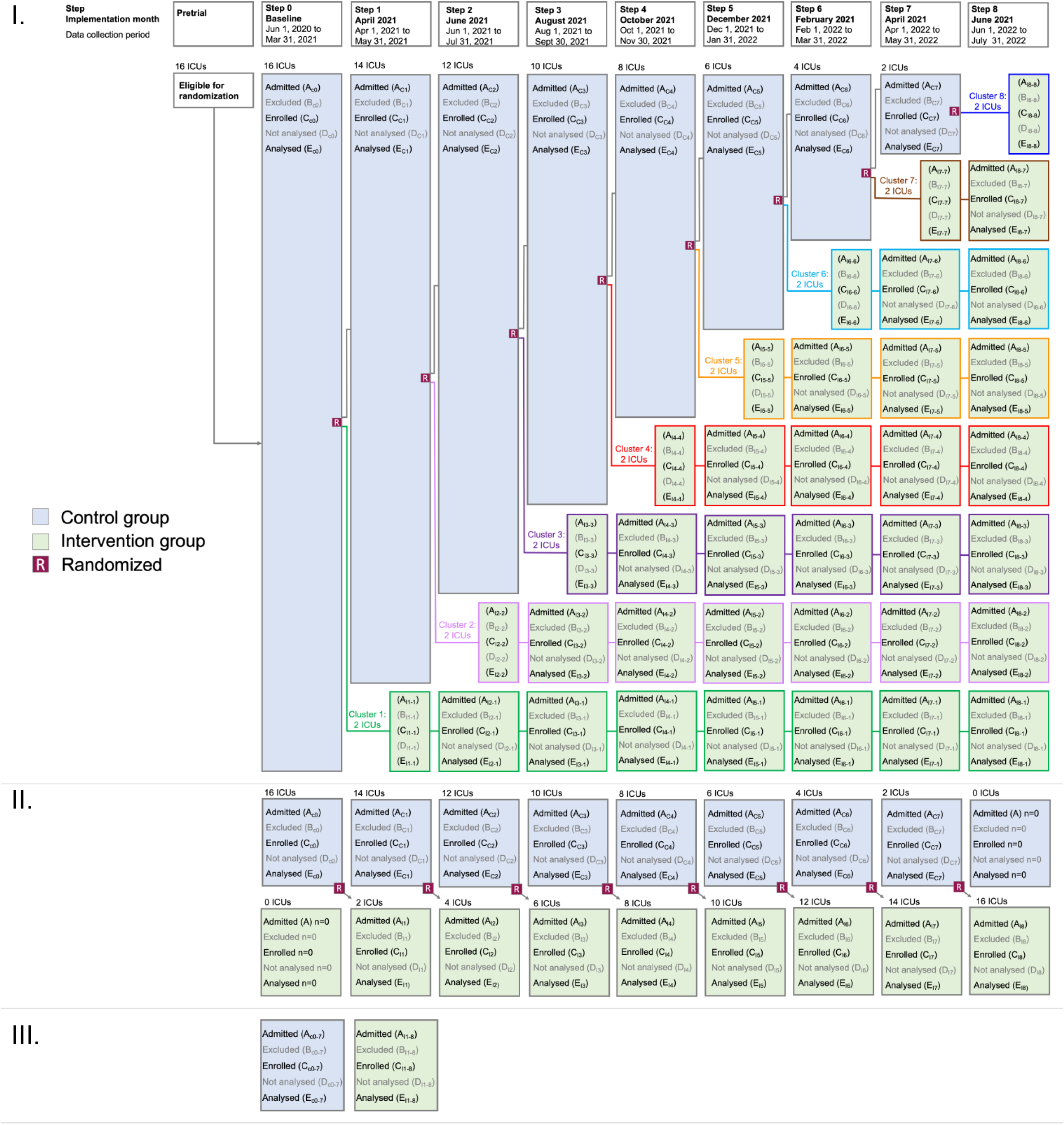
CONSORT Diagram I=patients by step. II=patients by cluster. III=patients at trial level. Admitted (A) = The number of patients admitted. Excluded (B) = The number of patients who were excluded e.g. NOT mechanically ventilated. Enrolled (C) = the number of patients who were enrolled. (C=A - B). Not analysed (D) = the number of patients who were NOT analysed e.g. no chart available. Analysed (E) = the number of patients who were analyzed (E=C - D). _c_=Control_. I_=Intervention. In each box the symbols represent the analysis status (A, B, C, D, E), whether it is control (c) or intervention (I) group, step of the stepped wedge (0 to 8), and in section I, for the intervention groups, which cluster (1-8). For example, AI5-2 is the number of patients admitted (A) in the intervention group (_I_) in the step 5 (_5_) of the study period for cluster 2 sites (_-2_).

### 7.4 Withdrawal / Follow-up

At the site level – a site will be withdrawn from the study if they decline or refuse to implement the pathway. All sites agreed to participate.

At the patient level, in the case of a patient transfer from a site where implementation is active to a non-active site or vice-versa, a patient will be deemed to be assigned to that original site for the purposes of assessing all outcomes if they have spent 48 hours or longer mechanically ventilated at that original site. This is based on a previous study that demonstrated that a lung protective ventilation strategy was most influential within the first 48 hours of initiation of invasive mechanical ventilation following ARDS diagnosis.(19) A sensitivity analysis will be conducted using alternative thresholds of 24 hours, 72 hours, and 1 week to test the robustness of this finding (see Section 8.2.1).

We will present any loss to follow-up but we expect this to be minimal due to electronic data collection.

### 7.5 aseline patient characteristics

Categorical data will be summarized by frequencies and percentages. Continuous data will be summarized as medians and IQR. Tests of statistical significance will not be undertaken for baseline characteristics; rather the clinical importance of any imbalance will be noted. Appendix Table 2 lists the baseline patient characteristics and how they will be reported.

## 8 Analysis

### 8.1 Outcome measures

#### 8.1.1 Primary clinical effectiveness outcome

The primary clinical outcome is **28-day ventilator free days (VFDs)** (in-hospital) a composite outcome of survival and days spent not ventilated over the first 28 days. 28-day VFDs are measured per admission, censored at hospital discharge and reported as mean (SD) and median (IQR).

#### 8.1.2 Primary implementation outcome

The primary implementation outcome is a **composite fidelity score (CFS)** that awards points for up to 5 key fidelity indicators that are met and is reported as a percentage. The individual indicators are measured per admission (height) or daily. Trends in mean CFS by time since implementation will be presented for all mechanically ventilated patients, patients with HRF, and patients with ARDS (definition 2, see Appendix Table 1 for definition).

#### 8.1.3 Secondary clinical effectiveness outcomes

1. *28-day hospital, ICU, and hospital survival* are measured per admission and reported as frequency with proportion of patients. 28-day hospital survival is measured at 28-days and censored at hospital discharge. Hospital survival is censored at 90 days. The first day of ventilation is day 0. 28-day hospital survival is the component of VFDs that reflects survival.
2. *Ventilator duration* is the number of ventilated days. If a patient is invasively mechanically ventilated via endotracheal tube or tracheostomy for any period of time in a 24-hour period (0000-2359) this is considered a ventilated day. A ventilated day is the component of VFDs that reflects duration of ventilation.
3. *Driving Pressure*. Driving pressure is calculated on patients ventilated with PF ratio (partial pressures of oxygen (PaO2) / fraction of inspired oxygen (FiO2)) ≤ 300 on a controlled mode as plateau pressure – positive end expiratory pressure (PEEP). It is reported throughout the ICU stay as median (IQR).
4. *Mechanical power*. Mechanical power is calculated on patients ventilated with PF ratio ≤ 300 on a controlled mode using the formula *Power* = *0.098*respiratory rate•(tidal volume/1000)*(Peak Pressure* – (0.5 *• Driving Pressure))*.(20) It is reported throughout the ICU stay as median (IQR).
5. *Length of Stay (LOS)*. ICU and hospital LOS are measured per admission and reported as median (IQR). Hospital LOS is censored at 90 days.
6. *Utilization of veno-venous Extracorporeal Membrane Oxygenation (VV-ECMO)*. Utilization of VV-ECMO is measured per admission and reported as frequency with proportion of patients.

#### 8.1.4 Secondary implementation outcomes

##### Fidelity

Secondary fidelity outcomes are process of care indicators that reflect the five key steps of the pathway:

1. Proportion of patients ventilated with a *height ever documented (*step 1) are measured per admission and reported as frequency (proportion). Additional secondary height outcomes are detailed in Appendix Table 3.
2. The proportion of eligible patient days (PF ratio≤ 300) who receive a *tidal volume ≤ 8ml/kg* predicted body weight on controlled mode ventilation (step2/3) is measured daily and reported as a frequency (proportion). If height is not documented, a tidal volume indicator is determined based on using an average height of:
  - 162cm for females [Predicted Body Weight 54.2kg, tidal volume <=434ml]
  - 176cm for males [Predicted Body Weight 71.5kg, tidal volume <=572ml]
  - Predicted body weight will be calculated as previously described.(21) If inhaled or set tidal volume is not available, exhaled tidal volume is used.
3. The proportion of eligible patient days (PF ratio ≤ 300) who have a *plateau pressure measured* on controlled mode ventilation (step 3) is measured daily and reported as a frequency (proportion).
4. The proportion of eligible patient days (PF ratio ≤ 150) *receiving neuromuscular blockade* on controlled mode ventilation (step 4) is measured daily and reported as a frequency (proportion).
5. The proportion of eligible patient days (PF ratio ≤ 150 and FiO2 ≥ 0.60) *receiving prone ventilation* on controlled mode ventilation (step 5) is measured daily and reported as a frequency (proportion).

Please see Appendix Table 3 for full details on the criteria for eligible patient days and definitions for each process of care indicator.

##### Acceptability

The acceptability outcomes assess clinician perceptions about the pathway and are based on the seven component constructs of the Theoretical Framework of Acceptability (TFA) listed below.(22) These are measured on a five-point Likert scale, a median of four or above indicates agreement.

1. *Composite acceptability score* is the proportion of the seven TFA constructs (see below 2 to 8) on the acceptability survey graded with a median score of four or above on a five-point Likert scale, indicating agreement.
2. *Intervention coherence* (the extent to which the clinician understands the intervention).
3. *Opportunity costs* (benefits or costs to the clinician for using the pathway).
4. *Perceived effectiveness* of the pathway (the extent to which the intervention is perceived by clinicians as likely to achieve its purpose).
5. *Self-efficacy* (a clinician’s confidence that they can use the pathway).
6. *Affective attitude* (how a clinician feels about the intervention).
7. *Burden* (a clinician’s perceived amount of effort required to participate in the intervention).
8. *Ethicality* (the extent to which the intervention aligns with a clinician’s value system).

See Appendix Table 3 for additional details of outcomes.

### 8.2 Analysis Methods

#### 8.2.1 Clinical effectiveness

Clinical outcomes will be analyzed at the patient-level. For the primary analysis, we will compare the mean 28-day VFDs pre-implementation and post-implementation using a mixed effects linear regression model to account for clustering of patients within site. Secondary clinical outcomes will be similarly compared pre-implementation and post-implementation using mixed effects linear or logistic regression models, as appropriate. All models will be adjusted for age, sex, severity of illness (sequential organ failure assessment score on admission) and severity of hypoxemia on admission based on PF ratio, as well as type and size of ICU. We will include time (days) in the models to account for secular trends over time, since failure to include such time effects can bias estimates of effect sizes. Data from the 1-month implementation transition phase within each step will not be included in the analysis of primary and secondary outcomes. If the distribution of a continuous outcome is skewed, a log-transformation of the outcome will be considered if applicable. A two-sided p-value < 0.05 will indicate statistical significance.

##### Sensitivity analysis

As a sensitivity analyses, we will analyze VFDs using a time-to-event analysis censored at 28 days using Fine and Gray competing risk regression since we have two mutually exclusive potential endpoints (successful extubation or death). If the proportional hazards assumption is not satisfied, the subdistribution hazard ratio obtained from the Fine and Gray model can be interpreted as the average subdistribution hazard ratio.(18) Schoenfeld-type residuals will be used to assess the proportional subdistribution hazard assumption.(23, 24) Differences in secondary outcomes pre and post-implementation will be analyzed using mixed effects linear and logistic regression models accounting for clustering of patients within site, as appropriate. For VFDs, we will also conduct a sensitivity analysis in which we exclude patients cared for in non-traditional ICU settings due to potential differences with patient cared for in traditional ICU settings (electronic data extraction). In the case of transfer delays out of the ICU due to bed availability, a sensitivity analysis will be conducted on ICU LOS. The sensitivity analysis will be conducted on ICU LOS by excluding ICU avoidable days and instead use the date ready for transfer.

##### 8.2.1.1 Subgroup analysis

The following subgroup analyses will be conducted for both the primary effectiveness outcome (28d VFDs) and also the primary implementation outcome (CFS). We will test for heterogeneity of treatment effect across these subgroups and report the corresponding p-value for interaction with a p-value less than 0.05 being deemed significant. To account for multiple testing for the subgroup analyses, we will report the false discovery rate.

- High vs low ICU volume (split at the median, over study period)
  ∘ *Low volume ICUs most likely to improve VFDs given lower baseline CFS*
- HRF (HRF vs non-HRF)
  ∘ *HRF patients most likely to improve VFDs as eligible to get more elements of pathway*
- ARDS definition 2 (see Appendix 1, Table 1 & 4) (ARDS vs non-ARDS)
  ∘ *ARDS patients most likely to improve VFDs as eligible to get more elements of pathway*
- Females vs males
  ∘ *Females most likely to improve VFDs given lower baseline CFS*
- Covid positive vs Covid negative
  ∘ *COVID patients most likely to improve VFDs as eligible to get more elements of pathway*
- Cardiac Surgery vs non-cardiac surgery patients
  ∘ *Non cardiac surgery patients most likely to improve VFDs as eligible to get more elements of pathway*
- Average height of patients (3 categories: quartile 1, quartile 2 and 3, quartile 4)
  ∘ *Lower quartile height patients to most likely to improve VFDs given lower baseline lung protective strategies*
- Severity of HRF within the first 24 hours of mechanical ventilation (severe vs moderate vs mild)
  ∘ *Severe HRF patients most likely to improve VFDs as eligible to get more elements of pathway*
- Age >60 vs 60 and under (median age)
  ∘ *Age > 60 patients most likely to improve VFDs as mortality at presentation is higher*
- Weight by Body Mass Index classifications (<18.5, 18.5 to <25, 25 to <30, >30)
  ∘ *Higher BMI patients to most likely to improve VFDs given lower baseline lung protective strategies*
- Severity of illness high vs low SOFA score (SOFA score <12 vs 12 or more)
  ∘ *SOFA > 12 patients most likely to improve VFDs as mortality at presentation is higher*

#### 8.2.2 Implementation

##### Fidelity

Quantitative assessment of fidelity will be tracked using process of care indicators that reflect the five key steps of the pathway. Differences in fidelity outcomes pre-implementation and post-implementation will be analyzed similarly to the effectiveness clinical outcomes using mixed effects regression models.

##### Acceptability (surveys)

Survey data will be presented as aggregated frequencies with proportions. Data will be stratified by clinician profession, years of experience, and type of institution. Differences will be compared using Fisher’s exact test or Chi-squared test for categorical variables, or the Wilcoxon rank-sum test or Kruskal Wallis test for Likert scale data, as appropriate.

### 8.3 Missing Data

Outcome data is expected to be available for all ventilated ICU patients admitted to the study ICUs during the study period as all clinical effectiveness and patient characteristics data is available electronically and will be extracted retrospectively following study completion. If patient data is not available electronically, data will be extracted from paper charts where available. If a PF ratio is unavailable, for example due to an arterial blood gas not being obtained or a patient does not having an arterial line, a non-invasive approach using pulse oximetry and the peripheral oxygen saturation (SpO2:FiO2) ratio will be used as previously described.(25-27)

### 8.4 Additional Analysis

Additional statistical analysis is not currently required.

### 8.5 Harms

Safety reporting is not being done as the intervention is not experimental it is standard of care. Harms are assessed in outcomes of VFDs and survival.

### 8.6 Statistical software

R will be used to carry out analysis.

### 8.7 References

References for statistical methods are listed below. Data management is detailed in the Data Access, Transfer, Encryption, and Storage sections of the protocol. The *Trial Master File* and *Statistical Master*

File are separate files and stored on a secure password-protected AHS computer held by the study biostatistician with restricted access.

## Data Availability

This is a statistical analysis plan. At the time of this dissemination no retrieval of electronic data or analyses has been conducted.

## 9 Additional Information

### 9.1 Health Economics

Details of the health economics analysis will be outlined in a separate Health Economics Analysis Plan. In this study we consider LOS both a clinical effectiveness and economic outcome.

### 9.2 Scientific Steering Group

The Protocol and Statistical Analysis plan have been reviewed by the TheraPPP Scientific Steering Group (see Appendix Table 5).

## 2 Abbreviations

ABG: Arterial Blood Gas
ARDS: Acute Respiratory Distress Syndrome
CXR: Chest x-ray
CFS: Composite Fidelity Score
CI: Confidence Interval
FiO_2_: Fraction of Inspired Oxygen
HRF: Hypoxemic Respiratory Failure
ICU: Intensive Care Unit
ICC: Intraclass Correlation Coefficient
IQR: Interquartile range
LOS: Length of Stay
PaO2: Partial pressure of oxygen
PF: ratio PaO2/FiO2
PEEP: Positive End Expiratory Pressure
PBW: Predicted Body Weight
SD: Standard Deviation
SpO2: Peripheral oxygen saturation
TFA: Theoretical Framework of Acceptability
VFDs: Ventilator Free Days
TV: Tidal volume
VV-ECMO: Veno-venous Extracorporeal Membrane Oxygenation

## 10 Signatures of approval

This SAP version 1.0 February 22, 2022 has been reviewed in detail by the Venting Wisely Scientific Steering Group and approved for dissemination and release. At the time of this dissemination no retrieval of electronic data or analyses has been conducted.

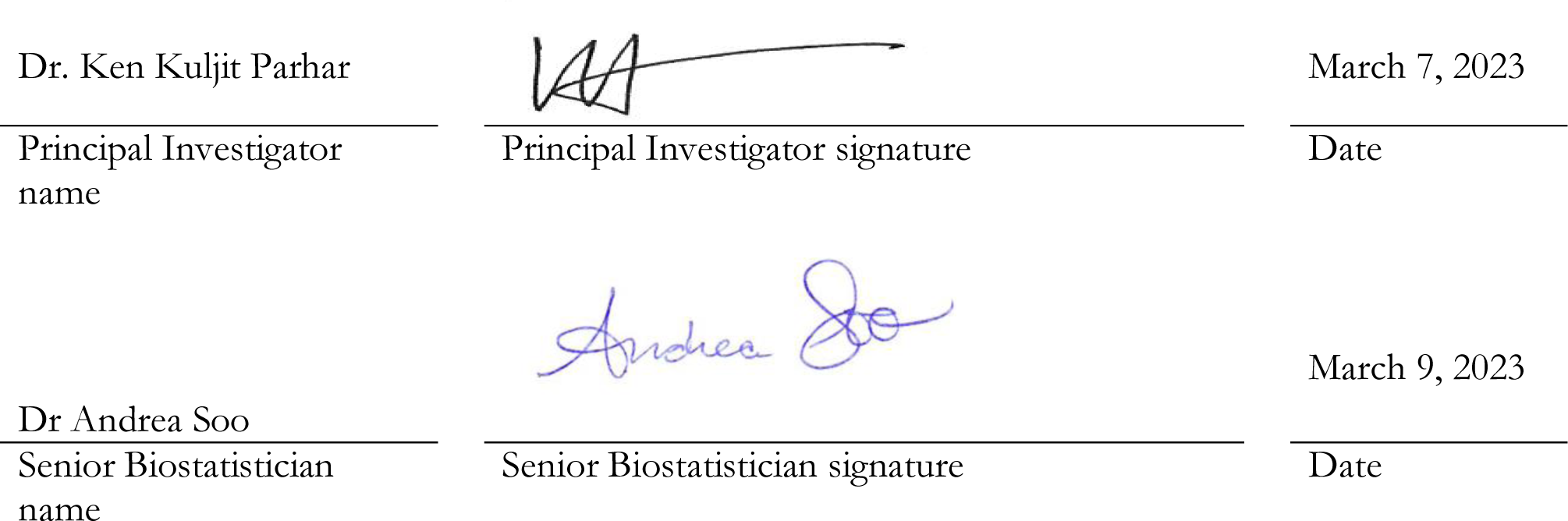

