## Supplemental Tables for "Statistical analysis plan for the Identification and Treatment of Hypoxemic Respiratory Failure (HRF) and ARDS with Protection, Paralysis, and Proning: a type-1 hybrid stepped-wedge cluster randomized effectiveness-implementation study"

### Appendix

**Table 1.** Criteria for definitions of Hypoxemic Respiratory Failure and ARDS

| Definition | PF ratio | Timing of ABG | CXR | Dx |
|---|---|---|---|---|
| HRF | ≤300 | At least 1 PF ratio 0000-0800 | N/A | N/A |
| Sustained HRF | ≤300 | At least 1 PF ratio on any ABG 6-36 hours after initial HRF diagnosis (2 ABGs) | N/A | N/A |
| Sustained ARDS | ≤300 | Any ABG done 6-36 hours after initial HRF diagnosis (2 ABGs) | Bilateral infiltrates | All **excluding** CHF & cardiogenic shock |
| eCritical ARDS **definition 1** (**exclusion** method) | ≤300 | Any ABG done 6-36 hours after initial HRF diagnosis (2 ABGs) | * | **Excludes** all primary diagnoses (e.g., cardiac conditions) not consistent with ARDS in eCritical† |
| eCritical ARDS **definition 2** (**inclusion** method) | ≤300 | Any ABG done 6-36 hours after initial HRF diagnosis (2 ABGs) | ** | **Includes** only conditions that could be indirect and direct causes of ARDS† |

ABG=arterial blood gas. ARDS=acute respiratory distress syndrome. CHF=congestive heart failure. CXR=chest x-ray. Dx=diagnoses. eCritical=data repository of patient-specific critical care clinical information. HRF=hypoxemic respiratory failure. PF ratio=PaO2/FiO2.

*When comparing the performance of the **exclusion** method definition to one that requires the **addition** of bilateral infiltrates on CXR, the specificity was 89.6% (95% CI 87.1%-91.8%) while the sensitivity was 100% (95% CI 98.3%-100%) with a kappa of 0.74 (95% CI 0.70-0.78).
**When comparing the performance of this definition to one that requires the **addition** of bilateral infiltrates on CXR, the specificity was 82.4% (95% CI 79.2%-85.3%) while the sensitivity was 100% (95% CI 98.7%-100%) with a kappa of 0.80 (95% CI 0.76-0.85).

†See Appendix Table 4 for a complete list of conditions defined as ARDS and non-ARDS by the exclusion and inclusion methods (ARDS definition 1 & 2).

**Table 2.** List of baseline characteristics of mechanically ventilated patients to be summarized

| Patient characteristic | Reported as |
|---|---|
| Age | median (IQR) |
| Sex | n (%) |
| Weight | median (IQR) |
| Height | median (IQR) |
| Predicted body weight | median (IQR) |
| Any comorbidity | n (%) |
| AIDS | |
| Acute renal failure | |
| Congestive heart failure | |
| Cirrhosis | |
| Dialysis | |
| Diabetes | |
| Hepatic failure | |
| Immunosuppression | |
| Leukemia | |
| Lymphoma | |
| Metastatic Cancer | |
| Respiratory insufficiency | |
| SOFA score on admission | median (IQR) |
| APACHE II score on admission | median (IQR) |
| Clinical Frailty Score | median (IQR) |
| Admission type | n (%) |
| Elective | |
| Emergent | |
| No surgery | |
| Unknown | |
| Reason for ICU admission | n (%) |
| Medical | |
| Surgical | |
| Neurological | |
| Trauma | |
| Unknown | |
| First PF ratio closest to 0800 | median (IQR) |
| Median PF ratio within the first 24 hours of being ventilated$^{\pm}$ | median (IQR) |
| Tidal volume (exhaled) (mL/kg)$^{\pm}$ | median (IQR) |
| Tidal volume (exhaled) (mL/kg) on controlled mode$^{\pm}$ | median (IQR) |
| Respiratory rate$^{\pm}$ | median (IQR) |
| Minute ventilation$^{\pm}$ | median (IQR) |
| PEEP$^{\pm}$ | median (IQR) |
| Plateau pressure$^{\pm}$ | median (IQR) |
| Plateau pressure on controlled mode$^{\pm}$ | median (IQR) |
| Peak pressure$^{\pm}$ | median (IQR) |
| Mean airway pressure$^{\pm}$ | median (IQR) |
| Driving pressure on controlled mode$^{\pm}$ | median (IQR) |
| Mechanical power on controlled mode$^{\pm}$ | median (IQR) |
| Hypoxemic respiratory failure (HRF)* | n (%) |
| ARDS (definition 2)** | n (%) |
| Vasoactive medications | n (%) |
| Continuous renal replacement therapy | n (%) |
| Intermittent hemodialysis | n (%) |

$^{\pm}$Within the first 24 hours of being ventilated. *Based on having at least 1 PF ≤300 on an arterial blook gas (ABG) done between 0000-0800. **HRF sustained (Based on PF ≤ 300 on any ABG done 6-36 hours after initial HRF diagnosis) (2 ABGs) + includes only conditions that could be direct and indirect causes of ARDS. APACHE=Acute Physiology and Chronic Health Evaluation. ARDS=acute respiratory distress syndrome. Driving pressure=Plateau pressure - Positive End Expiratory Pressure. IQR =interquartile range. PEEP=Positive End Expiratory Pressure. PF ratio=$PaO_2$:$FiO_2$ ratio. Mechanical power=0.098*resp rate*(tidal volume/1000)*(peak-(0.5*driving)). SOFA=Sequential Organ Failure Assessment. VFD=ventilator free days.

Venting Wisely
HRF & ARDS Pathway

**Table 3.** Primary and secondary effectiveness, fidelity, and acceptability outcomes

| **Clinical Effectiveness Outcomes** | **Patient or subgroup** | **Timepoints at which the outcomes are measured** | **Primary or Secondary Outcome** | **Reporting of results (unit of measurement)** |
|---|---|---|---|---|
| 28-day ventilator-free days (VFDs)<br>(a composite of survival & days spent not ventilated over the first 28 days) | All pts / subgroups | Per admission 28-days<br>Censored at hospital discharge | Primary | Mean (SD) and Median with interquartile range (IQR) |
| ICU survival | All pts / select subgroups€ | Per admission | Secondary | Frequency with proportion of pts |
| ICU Length of Stay | All pts / select subgroups€ | Per admission | Secondary | Median with IQR |
| 28-day hospital survival | All pts / select subgroups€ | Per admission 28-days<br>The first day of MV is day 0<br>Censored at hospital d/c | Secondary | Frequency with proportion of pts |
| Ventilator duration | All pts /select subgroups€ | Per admission | Secondary | Median with IQR |
| Hospital survival | All pts /select subgroups€ | Per admission<br>Censored 90 days after MV | Secondary | Frequency with proportion of pts |
| Hospital Length of stay | All pts /select subgroups€ | Per admission<br>Censored 90 days after MV | Secondary | Median with IQR |
| Driving Pressure<br>(Plateau pressure – PEEP) | Pts MV with PF ratio ≤300 on controlled mode€ | Throughout the ICU stay | Secondary | Median with IQR |
| Mechanical Power<br>Calculated using the formula:<br>Power = 0.098*respiratory rate•*(tidal volume/1000)*(Peak Pressure – (0.5 • Driving Pressure) | Pts MV with PF ratio ≤300 on controlled mode€ | Throughout the ICU stay | Secondary | Median with IQR |
| Utilization of veno-venous Extracorporeal Membrane Oxygenation (VV-ECMO) | All pts /select subgroups€ | Per admission | Secondary | Frequency with proportion of pts |
| **Implementation outcomes - Fidelity Indicators** | **Patient or subgroup** | **Timepoints at which the outcomes are measured** | **Primary or Secondary Outcome** | **Reporting of results (unit of measurement)** |
| Composite fidelity score (CFS)<br>The CFS awards points for fidelity indicators. It can be interpreted as the average proportion of time pathway elements are appropriately performed, it:<br>- Includes all MV patients<br>- Is based on pts being eligible for up to 4 possible interventions per day and height ever documented during the ICU stay (specific indicators indicated by asterisk* below)<br>- A day is only included if pt is eligible for that day<br>- Height measurement only contributes 1 point to the score if it is ever measured during the ICU stay | All pts / subgroups<br>Pt subgroups for individual fidelity indicators are indicated by asterisk* in rows below | Per admission (height) & daily (tidal volume, plateau pressure, neuromuscular blockade and proning) | Primary | For each pt, the CFS is calculated as the number of times pathway elements were appropriately performed out of the number of times the pt was eligible for pathway elements throughout their ICU stay. The CFS is reported as Median (IQR) % and Mean (SD) % |
| *Height ever documented | MV pts | Per admission | Secondary | Frequency with proportion of pts |
| Height documented within 1 hour of admission to ICU | MV pts | Per admission | Secondary | Frequency with proportion of pts |
| Height documented within 2 hours of admission to ICU | MV pts | Per admission | Secondary | Frequency with proportion of pts |
| Time in minutes to height measurement from ICU admission (among pts with height ever documented) | MV pts with a height measured | Per admission | Secondary | Median with IQR |
| ***Tidal volume (TV) ≤ 8ml/kg PBW: Previously noted as *Days of safe ventilation*<br>If height is not documented, TV indicator is determined based on using an average height of:<br>• 162cm for females [PBW 54.2kg, TV <=434ml]<br>• 176cm for males [PBW 71.5kg, TV <=572ml] | MV pts:<br>- ABG done that day between 0000-0800<br>- PF ratio ≤ 300 on that day<br>- On controlled mode | Daily | Secondary | Median (IQR) and mean (SD) proportion of eligible days<br>Outcome is assessed QD, but then summarized as a proportion for each pt, and then the median or mean is taken |

Venting Wisely HRF & ARDS Pathway

| | | | | |
|---|---|---|---|---|
| TV set will be used for volume-controlled mode and TV inhaled will be used for pressure-controlled mode. If inhaled or set TV is not available, exhaled TV is used. Indicator is based on the median daily TV being ≤ 8 ml/kg PBW | | | | |
| *****Plateau pressure measured | *Patients ventilated<br>- ABG done that day<br>- PF ratio ≤ 300 on that day<br>- On controlled mode | Daily | Secondary | " |
| *****Receive any neuromuscular blockade in the consider group (including as little as a single bolus to an infusion) | *Patients ventilated<br>- ABG done that day<br>- PF ratio ≤ 150 on that day<br>- On controlled mode | Daily | Secondary | " |
| *****Patient proned for those in the consider group | *Patients ventilated<br>- ABG done that day<br>- PF ratio ≤150 and FiO2 ≥0.60 on that day<br>- On controlled mode<br>- Not receiving ECMO that day | Daily | Secondary | " |
| **Implementation Outcomes – Acceptability (survey)** | **Patient or subgroup** | **Timepoints at which the outcomes are measured** | **Primary or Secondary Outcome** | **Reporting of results (unit of measurement)** |
| C**omposite Acceptability Score:** Summary of pathway acceptability measured using the 7 Theoretical Framework of Acceptability (TFA) component constructs (listed below†) | Survey clinicians and pathway educators / champions | Two to six months after implementation | Secondary | Proportion of TFA components with median score of 4 or 5 on a 5-point Likert scale, indicating agreement |
| **†Affective attitude** (How an individual feels about the intervention) | " | " | Secondary | Median (IQR) |
| **†Burden** (The perceived amount of effort that is required to participate in the intervention) | " | " | Secondary | " |
| **†Ethicality** (The extent to which the intervention has a good fit with an individual's value) | " | " | Secondary | " |
| **†Intervention coherence** (The extent to which the participant understands the invention and how it works) | " | " | Secondary | " |
| **†Opportunity costs** (The extent to which benefits, profits, or values must be given up to engage in the intervention | " | " | Secondary | " |
| **†Perceived effectiveness** (The extent to which the intervention is perceived as likely to achieve its purpose) | " | " | Secondary | " |
| **†Self-efficacy** (The participant's confidence that they can perform the behavior(s) required to participate in the intervention) | " | " | Secondary | " |

€hypoxemic respiratory failure and ARDS subgroups only. ABG=arterial blood gas. CFS=composite fidelity score. d/c=discharge. ECMO=extra corporeal membrane oxygenation. ICU=intensive care unit. IQR= Interquartile range. MV=mechanical ventilation. PEEP=positive end expiratory unit. PF ratio=PaO2/FiO2. PBW=predicted body weight. QD=daily. SD=standard deviation. Pts=patients. TFA=theoretical framework of acceptability. TV=tidal volume. VFDs=ventilator free days. VV-ECMO= veno-venous Extracorporeal Membrane Oxygenation.

**Table 4.** Conditions defined as ARDS and non-ARDS by Definition 1 (Exclusion method) and Definition 2 (Inclusion method)

| Condition | Definition 1 | Definition 2 |
|---|---|---|
| Abdomen only trauma | ARDS | ARDS |
| Abdomen only trauma, surgery for | ARDS | ARDS |
| Abdomen/extremity trauma | ARDS | ARDS |
| Abdomen/extremity trauma, surgery for | ARDS | ARDS |
| Abdomen/face trauma | ARDS | ARDS |
| Abdomen/face trauma, surgery for | ARDS | ARDS |
| Abdomen/multiple trauma | ARDS | ARDS |
| Abdomen/multiple trauma, surgery for | ARDS | ARDS |
| Abdomen/pelvis trauma | ARDS | ARDS |
| Abdomen/pelvis trauma, surgery for | ARDS | ARDS |
| Abdomen/spinal trauma | ARDS | ARDS |
| Abdomen/spinal trauma, surgery for | ARDS | ARDS |
| Ablation or mapping of cardiac conduction pathway | nonARDS | nonARDS |
| Abscess, neurologic | ARDS | nonARDS |
| Abscess/infection-cranial, surgery for | ARDS | nonARDS |
| Acid-base electrolyte disturbance | ARDS | nonARDS |
| Addisons disease | ARDS | nonARDS |
| Adrenal neoplasm (including pheochromocytoma) | ARDS | nonARDS |
| Adrenalectomy | ARDS | nonARDS |
| Alcohol withdrawal | ARDS | nonARDS |
| All Burn Patients | ARDS | ARDS |
| All Surgical Burn Patients | ARDS | ARDS |
| Amputation (non-traumatic) | ARDS | nonARDS |
| Amyotrophic lateral sclerosis | ARDS | nonARDS |
| Anaphylaxis | ARDS | nonARDS |
| Anastomosis, vascular | ARDS | nonARDS |
| Anemia | ARDS | nonARDS |
| Aneurysm repair, ventricular | ARDS | nonARDS |
| Aneurysm, abdominal aortic | ARDS | nonARDS |
| Aneurysm, abdominal aortic; with dissection | ARDS | nonARDS |
| Aneurysm, abdominal aortic; with rupture | ARDS | nonARDS |
| Aneurysm, dissecting aortic | ARDS | nonARDS |
| Aneurysm, thoracic aortic | ARDS | nonARDS |
| Aneurysm, thoracic aortic; with dissection | ARDS | nonARDS |
| Aneurysm, thoracic aortic; with rupture | ARDS | nonARDS |
| Aneurysm/pseudoaneurysm, other | ARDS | nonARDS |
| Aneurysms, repair of other (except ventricular) | ARDS | nonARDS |
| Angina, stable (asymp or stable pattern of symptoms w/meds) | nonARDS | nonARDS |
| Angina, unstable (angina interferes w/quality of life or meds are tolerated poorly) | nonARDS | nonARDS |
| Aortic and Mitral valve replacement | nonARDS | nonARDS |
| Aortic valve replacement (isolated) | nonARDS | nonARDS |
| Apnea-sleep; surgery for (i.e. UPPP -uvulopalatopharyngoplasty) | ARDS | nonARDS |
| Apnea, sleep | ARDS | nonARDS |
| Appendectomy | ARDS | nonARDS |
| ARDS-adult respiratory distress syndrome, non-cardiogenic pulmonary edema | ARDS | ARDS |
| Arrest, respiratory (without cardiac arrest) | ARDS | nonARDS |
| Arteriovenous malformation, surgery for | ARDS | nonARDS |
| Arthritis, septic | ARDS | nonARDS |
| Asthma | ARDS | nonARDS |
| Atelectasis | nonARDS | nonARDS |
| Atrial Septal Defect (ASD) Repair | nonARDS | nonARDS |
| Biopsy, brain | ARDS | nonARDS |
| Biopsy, open lung | ARDS | nonARDS |
| Bladder repair for perforation/rupture | ARDS | nonARDS |
| Bleeding-lower GI, surgery for | ARDS | nonARDS |
| Bleeding-other GI, surgery for | ARDS | nonARDS |
| Bleeding-upper GI, surgery for | ARDS | nonARDS |
| Bleeding, GI-location unknown | ARDS | nonARDS |
| Bleeding, GI from esophageal varices/portal hypertension | ARDS | nonARDS |
| Bleeding, lower GI | ARDS | nonARDS |
| Bleeding, upper GI | ARDS | nonARDS |
| Blood transfusion reaction | ARDS | nonARDS |
| Bone marrow transplant | ARDS | ARDS |
| Bullectomy | ARDS | nonARDS |
| Burr hole placement | ARDS | nonARDS |
| CABG alone, coronary artery bypass grafting | nonARDS | nonARDS |
| CABG alone, redo | nonARDS | nonARDS |
| CABG redo with other operation | nonARDS | nonARDS |
| CABG redo with valve repair/replacement | nonARDS | nonARDS |

| | | |
|---|---|---|
| CABG with aortic valve replacement | nonARDS | nonARDS |
| CABG with double valve repair/replacement | nonARDS | nonARDS |
| CABG with mitral valve repair | nonARDS | nonARDS |
| CABG with mitral valve replacement | nonARDS | nonARDS |
| CABG with other operation | nonARDS | nonARDS |
| CABG with pulmonic or tricuspid valve repair or replacement ONLY. | nonARDS | nonARDS |
| Cancer-colon/rectal, surgery for (including abdominoperineal resections) | ARDS | nonARDS |
| Cancer-esophageal, surgery for (abdominal approach) | ARDS | nonARDS |
| Cancer-laryngeal/tracheal, surgery for | ARDS | nonARDS |
| Cancer-other GI tract, surgery for (i.e. hepatoma, gallbladder etc.) | ARDS | nonARDS |
| Cancer-small intestinal, surgery for | ARDS | nonARDS |
| Cancer-stomach, surgery for | ARDS | nonARDS |
| Cancer oral/sinus, surgery for | ARDS | nonARDS |
| Cancer, colon/rectal | ARDS | nonARDS |
| Cancer, esophageal | ARDS | nonARDS |
| Cancer, laryngeal | ARDS | nonARDS |
| Cancer, lung | ARDS | nonARDS |
| Cancer, oral | ARDS | nonARDS |
| Cancer, other GI | ARDS | nonARDS |
| Cancer, pancreatic | ARDS | nonARDS |
| Cancer, stomach | ARDS | nonARDS |
| Cancer, tracheal | ARDS | nonARDS |
| CAPD catheter insertion | ARDS | nonARDS |
| Cardiac arrest (with or without respiratory arrest; for respiratory arrest see Respiratory System) | ARDS | nonARDS |
| Cardiomyopathy | nonARDS | nonARDS |
| Cardiovascular medical, other | ARDS | nonARDS |
| Cardiovascular surgery, other | nonARDS | nonARDS |
| Cellulitis and localized soft tissue infections | ARDS | nonARDS |
| Cellulitis and localized soft tissue infections, surgery for | ARDS | ARDS |
| Cerebrospinal fluid leak, surgery for | ARDS | nonARDS |
| Cesarean section | ARDS | nonARDS |
| Chest pain, epigastric | nonARDS | nonARDS |
| Chest/abdomen trauma | ARDS | ARDS |
| Chest/abdomen trauma, surgery for | ARDS | ARDS |
| Chest/extremity trauma | ARDS | ARDS |
| Chest/extremity trauma, surgery for | ARDS | ARDS |
| Chest/face trauma | ARDS | ARDS |
| Chest/face trauma, surgery for | ARDS | ARDS |
| Chest/multiple trauma | ARDS | ARDS |
| Chest/multiple trauma, surgery for | ARDS | ARDS |
| Chest/pelvis trauma | ARDS | ARDS |
| Chest/pelvis trauma, surgery for | ARDS | ARDS |
| Chest/spinal trauma | ARDS | ARDS |
| Chest/spinal trauma, surgery for | ARDS | ARDS |
| Chest/thorax only trauma | ARDS | ARDS |
| Chest/thorax only trauma, surgery for | ARDS | ARDS |
| CHF, congestive heart failure | nonARDS | nonARDS |
| Cholangitis | ARDS | nonARDS |
| Cholecystectomy/cholangitis, surgery for (gallbladder removal) | ARDS | nonARDS |
| Coagulopathy | ARDS | nonARDS |
| Coma/change in level of consciousness (for hepatic see GI, for diabetic see Endocrine, if related to cardiac arrest , see CV) | ARDS | nonARDS |
| Complications of prev. peripheral vasc. surgery,surgery for (i.e.ligation of bleeder, debridement, pseudoaneurysms, clots, fistula, etc.) | ARDS | nonARDS |
| Complications of previous GI surgery; surgery for (anastomotic leak, bleeding, abscess, infection, dehiscence, etc.) | ARDS | nonARDS |
| Complications of previous open-heart surgery, surgery for (i.e. bleeding, infection, mediastinal rewiring,leaking aortic graft etc.) | nonARDS | nonARDS |
| Complications of previous open heart surgery (i.e. bleeding, infection etc.) | nonARDS | nonARDS |
| Complications of previous spinal cord surgery, surgery for | ARDS | nonARDS |
| Congenital Defect Repair (Other) | ARDS | nonARDS |
| Connective tissue disease (mixed) | ARDS | nonARDS |
| Contusion, myocardial | ARDS | nonARDS |
| Cosmetic surgery (all) | ARDS | nonARDS |
| Cranioplasty and complications from previous craniotomies | ARDS | nonARDS |
| CVA, cerebrovascular accident/stroke | ARDS | nonARDS |
| Cyst, ruptured ovarian | ARDS | nonARDS |
| Cystectomy (other reasons) | ARDS | nonARDS |
| Cystectomy for neoplasm | ARDS | nonARDS |
| Defibrillator, automatic implantable cardiac; insertion of | nonARDS | nonARDS |
| Devices for spine fracture/dislocation | ARDS | nonARDS |
| Diabetic hyperglycemic hyperosmolar nonketotic coma (HHNC) | ARDS | nonARDS |
| Diabetic ketoacidosis | ARDS | nonARDS |
| Diverticular disease | ARDS | nonARDS |

| | | |
|---|---|---|
| Diverticular disease, surgery for | ARDS | nonARDS |
| Drug withdrawal | ARDS | nonARDS |
| Ectopic pregnancy (all) | ARDS | nonARDS |
| Efffusion, pericardial | nonARDS | nonARDS |
| Effusions, pleural | nonARDS | nonARDS |
| Embolectomy (with general anesthesia) | ARDS | nonARDS |
| Embolus, pulmonary | nonARDS | nonARDS |
| Emphysema/bronchitis | ARDS | nonARDS |
| Encephalitis | ARDS | nonARDS |
| Encephalopathies (excluding hepatic) | ARDS | nonARDS |
| Encephalopathy, hepatic | ARDS | nonARDS |
| Endarterectomy (other vessels) | ARDS | nonARDS |
| Endarterectomy, carotid | ARDS | nonARDS |
| Endocarditis | ARDS | nonARDS |
| Esophageal surgery, other | ARDS | nonARDS |
| Exenteration, pelvic-female | ARDS | nonARDS |
| Exenteration, pelvic -male | ARDS | nonARDS |
| Extremity only trauma | ARDS | ARDS |
| Extremity only trauma, surgery for | ARDS | ARDS |
| Extremity/face trauma | ARDS | ARDS |
| Extremity/face trauma, surgery for | ARDS | ARDS |
| Extremity/multiple trauma | ARDS | ARDS |
| Extremity/multiple trauma, surgery for | ARDS | ARDS |
| Face only trauma | ARDS | ARDS |
| Face only trauma, surgery for | ARDS | ARDS |
| Face/multiple trauma | ARDS | ARDS |
| Face/multiple trauma, surgery for | ARDS | ARDS |
| Facial surgery (if related to trauma, see Trauma) | ARDS | nonARDS |
| Fistula/abscess, surgery for (not inflammatory bowel disease) | ARDS | nonARDS |
| Fracture-pathological, non-union, non-traumatic, for fractures due to trauma see Trauma | ARDS | ARDS |
| Fusion-spinal/Harrington rods | ARDS | nonARDS |
| Gastrostomy | ARDS | nonARDS |
| Genitourinary medical, other | ARDS | nonARDS |
| Genitourinary surgery, other | ARDS | nonARDS |
| GI Abscess/cyst | ARDS | nonARDS |
| GI Abscess/cyst-primary, surgery for (for complications of GI surgery see below) | ARDS | nonARDS |
| GI medical, other | ARDS | nonARDS |
| GI Obstruction | ARDS | nonARDS |
| GI obstruction, surgery for (including lysis of adhesions) | ARDS | nonARDS |
| GI Perforation/rupture | ARDS | nonARDS |
| GI perforation/rupture, surgery for | ARDS | nonARDS |
| GI surgery, other | ARDS | nonARDS |
| GI Vascular insufficiency | ARDS | nonARDS |
| GI vascular ischemia, surgery for (resection) | ARDS | nonARDS |
| Graft for dialysis, insertion of | ARDS | nonARDS |
| Graft, aorto-femoral bypass | ARDS | nonARDS |
| Graft, aorto-iliac bypass | ARDS | nonARDS |
| Graft, femoral-femoral bypass | ARDS | nonARDS |
| Graft, femoral-popliteal bypass | ARDS | nonARDS |
| Grafting, skin (all) | ARDS | nonARDS |
| Grafts, all other bypass (except renal) | ARDS | nonARDS |
| Grafts, all renal bypass | ARDS | nonARDS |
| Grafts, removal of infected vascular | ARDS | nonARDS |
| Guillian-Barre syndrome | ARDS | nonARDS |
| Head (CNS) only trauma | ARDS | ARDS |
| Head (CNS) only trauma, surgery for | ARDS | ARDS |
| Head/abdomen trauma | ARDS | ARDS |
| Head/abdomen trauma, surgery for | ARDS | ARDS |
| Head/chest trauma | ARDS | ARDS |
| Head/chest trauma, surgery for | ARDS | ARDS |
| Head/extremity trauma | ARDS | ARDS |
| Head/extremity trauma, surgery for | ARDS | ARDS |
| Head/face trauma | ARDS | ARDS |
| Head/face trauma, surgery for | ARDS | ARDS |
| Head/multiple trauma | ARDS | ARDS |
| Head/multiple trauma, surgery for | ARDS | ARDS |
| Head/pelvis trauma | ARDS | ARDS |
| Head/pelvis trauma, surgery for | ARDS | ARDS |
| Head/spinal trauma | ARDS | ARDS |
| Head/spinal trauma, surgery for | ARDS | ARDS |

| | | |
|---|---|---|
| Heart-lung transplant | ARDS | nonARDS |
| Heart transplant | ARDS | nonARDS |
| Heat exhaustion/stroke | ARDS | nonARDS |
| Hematologic medical, other | ARDS | nonARDS |
| Hematologic surgery, other | ARDS | nonARDS |
| Hematoma, epidural | ARDS | nonARDS |
| Hematoma, epidural, surgery for | ARDS | nonARDS |
| Hematoma, subdural | ARDS | nonARDS |
| Hematoma, subdural, surgery for | ARDS | nonARDS |
| Hematomas | ARDS | nonARDS |
| Hemorrhage (for gastrointestinal bleeding GI-see GI system) (for trauma see Trauma) | ARDS | nonARDS |
| Hemorrhage, intra/retroperitoneal | ARDS | nonARDS |
| Hemorrhage, postpartum (female only) | ARDS | nonARDS |
| Hemorrhage/hematoma-intracranial, surgery for | ARDS | nonARDS |
| Hemorrhage/hematoma, intracranial | ARDS | nonARDS |
| Hemorrhage/hemoptysis, pulmonary | ARDS | nonARDS |
| Hemothorax | ARDS | nonARDS |
| Hepatic failure, acute | ARDS | nonARDS |
| Hepato-renal syndrome | ARDS | nonARDS |
| Hernia-hiatal, esophageal surgery for | ARDS | nonARDS |
| Herniorrhaphy | ARDS | nonARDS |
| Hip replacement, total (non-traumatic) | ARDS | nonARDS |
| Hydrocephalus, obstructive | ARDS | nonARDS |
| Hypertension-pulmonary, primary/idiopathic | ARDS | nonARDS |
| Hypertension, uncontrolled (for cerebrovascular accident-see Neurological System) | ARDS | nonARDS |
| Hyperthermia | ARDS | nonARDS |
| Hyperthyroid storm/crisis | ARDS | nonARDS |
| Hypoglycemia | ARDS | nonARDS |
| Hypothermia | ARDS | nonARDS |
| Hypothyroid/myxedema | ARDS | nonARDS |
| Hypovolemia (including dehydration. Do NOT include shock states.) | ARDS | nonARDS |
| Hysterectomy for cancer with or without lymph node dissection | ARDS | nonARDS |
| Hysterectomy for other benign neoplasm/fibroids | ARDS | nonARDS |
| Infarction, acute myocardial (MI) | nonARDS | nonARDS |
| Infection/abscess, other surgery for | ARDS | ARDS |
| Inflammatory bowel disease | ARDS | nonARDS |
| Inflammatory bowel disease, surgery for | ARDS | nonARDS |
| Kidney-pancreas transplant | ARDS | nonARDS |
| Kidney transplant | ARDS | nonARDS |
| Knee replacement, total (non-traumatic) | ARDS | nonARDS |
| Laminectomy/spinal cord decompression (excluding malignancies) | ARDS | nonARDS |
| Leukemia, acute lymphocytic | ARDS | nonARDS |
| Leukemia, acute myelocytic | ARDS | nonARDS |
| Leukemia, chronic lymphocytic | ARDS | nonARDS |
| Leukemia, chronic myelocytic | ARDS | nonARDS |
| Leukemia, other | ARDS | nonARDS |
| Liver-small bowel transplant | ARDS | nonARDS |
| Liver transplant | ARDS | nonARDS |
| Lung transplant, bilateral | ARDS | nonARDS |
| Lung transplant, single | ARDS | nonARDS |
| Lupus, systemic | ARDS | nonARDS |
| Lymph node dissection, pelvic or retroperitoneal (male) | ARDS | nonARDS |
| Lymphoma, Hodgkins | ARDS | nonARDS |
| Lymphoma, non-Hodgkins | ARDS | nonARDS |
| Lymphoma, non-Hodgkins; surgery for (including staging) | ARDS | nonARDS |
| Mastectomy (all) | ARDS | nonARDS |
| Meningitis | ARDS | nonARDS |
| Metabolic/endocrine medical, other | ARDS | nonARDS |
| Metabolic/endocrine surgery, other | ARDS | nonARDS |
| Mitral valve repair | nonARDS | nonARDS |
| Mitral valve replacement | nonARDS | nonARDS |
| Monitoring, hemodynamic (pre-operative evaluation) | ARDS | nonARDS |
| Musculoskeletal medical, other | ARDS | nonARDS |
| Myasthenia gravis | ARDS | nonARDS |
| Near drowning accident | ARDS | nonARDS |
| Neoplasm-cranial, surgery for (excluding transphenoidal) | ARDS | nonARDS |
| Neoplasm-spinal cord surgery or other related procedures | ARDS | nonARDS |
| Neoplasm, neurologic | ARDS | nonARDS |
| Nephrectomy (other reasons) | ARDS | nonARDS |
| Nephrectomy for neoplasm | ARDS | nonARDS |

| | | |
|---|---|---|
| Neurologic medical, other | ARDS | nonARDS |
| Neurologic surgery, other | ARDS | nonARDS |
| Neuromuscular medical, other | ARDS | nonARDS |
| Nontraumatic coma due to anoxia/ischemia | ARDS | nonARDS |
| Obesity-morbid, surgery for | ARDS | nonARDS |
| Obstruction-airway (i.e. acute epiglottitis, post-extubation edema, foreign body, etc.) | ARDS | nonARDS |
| Obstruction due to neoplasm ,surgery for; (with or without ileal-conduit) | ARDS | nonARDS |
| Obstruction due to nephrolithiasis, surgery for (with or without ileal-conduit) | ARDS | nonARDS |
| Obstruction/other, surgery for (with or without ileal-conduit) | ARDS | nonARDS |
| Oophorectomy with/without salpingectomy with/without lymph node dissection | ARDS | nonARDS |
| Orchiectomy with/without pelvic lymph node dissection | ARDS | nonARDS |
| Orthopedic surgery, other | ARDS | nonARDS |
| Overdose, alcohols (bethanol, methanol, ethylene glycol) | ARDS | nonARDS |
| Overdose, analgesic (aspirin, acetaminophen) | ARDS | nonARDS |
| Overdose, antidepressants (cyclic, lithium) | ARDS | nonARDS |
| Overdose, other toxin, poison or drug | ARDS | nonARDS |
| Overdose, sedatives, hypnotics, antipsychotics, benzodiazepines | ARDS | nonARDS |
| Overdose, street drugs (opiates, cocaine, amphetamine) | ARDS | nonARDS |
| Palsy, cranial nerve | ARDS | nonARDS |
| Pancreatitis | ARDS | ARDS |
| Pancreatitis, surgery for | ARDS | nonARDS |
| Pancytopenia | ARDS | nonARDS |
| Papillary muscle rupture | ARDS | nonARDS |
| Parathyroidectomy | ARDS | nonARDS |
| Pelvic relaxation (cystocele, rectocele, etc.) | ARDS | nonARDS |
| Pelvis/extremity trauma | ARDS | ARDS |
| Pelvis/extremity trauma, surgery for | ARDS | ARDS |
| Pelvis/face trauma | ARDS | ARDS |
| Pelvis/face trauma, surgery for | ARDS | ARDS |
| Pelvis/hip only trauma | ARDS | ARDS |
| Pelvis/hip only trauma, surgery for | ARDS | ARDS |
| Pelvis/multiple trauma | ARDS | ARDS |
| Pelvis/multiple trauma, surgery for | ARDS | ARDS |
| Pelvis/spinal trauma | ARDS | ARDS |
| Pelvis/spinal trauma, surgery for | ARDS | ARDS |
| Pericardial effusion/tamponade | ARDS | nonARDS |
| Pericardiectomy (total/subtotal) | ARDS | nonARDS |
| Pericarditis | nonARDS | nonARDS |
| Peritoneal lavage | ARDS | nonARDS |
| Peritonitis | ARDS | nonARDS |
| Peritonitis, surgery for | ARDS | nonARDS |
| Pneumonia, aspiration | ARDS | ARDS |
| Pneumonia, bacterial | ARDS | ARDS |
| Pneumonia, fungal | ARDS | nonARDS |
| Pneumonia, other | ARDS | ARDS |
| Pneumonia, parasitic (i.e. Pneumocystis pneumonia) | ARDS | nonARDS |
| Pneumonia, viral | ARDS | ARDS |
| Pneumothorax | nonARDS | nonARDS |
| Poisoning, carbon monoxide, arsenic, cyanide | ARDS | nonARDS |
| Pre-eclampsia/eclampsia (female only) | ARDS | nonARDS |
| Prostatectomy, suprapubic; for benign prostatic hypertrophy | ARDS | nonARDS |
| Prostatectomy, suprapubic; for cancer | ARDS | nonARDS |
| Pulmonary valve surgery | nonARDS | nonARDS |
| Renal bleeding | ARDS | nonARDS |
| Renal failure, acute | ARDS | nonARDS |
| Renal infection/abscess | ARDS | nonARDS |
| Renal neoplasm, cancer | ARDS | nonARDS |
| Renal obstruction | ARDS | nonARDS |
| Respiratory- medical, other | ARDS | nonARDS |
| Respiratory surgery, other | ARDS | nonARDS |
| Restrictive lung disease (i.e. sarcoidosis, pulmonary fibrosis) | ARDS | nonARDS |
| Rhabdomyolysis | ARDS | nonARDS |
| Rhythm disturbance (atrial, supraventricular) | nonARDS | nonARDS |
| Rhythm disturbance (conduction defect) | nonARDS | nonARDS |
| Rhythm disturbance (ventricular) | nonARDS | nonARDS |
| Scleroderma | ARDS | nonARDS |
| Seizures-intractable, surgery for | ARDS | nonARDS |
| Seizures (primary-no structural brain disease) | ARDS | nonARDS |
| Sepsis, cutaneous/soft tissue - other | ARDS | ARDS |
| Sepsis, GI - other | ARDS | ARDS |

| | | |
|---|---|---|
| Sepsis, GU/UTI (including bladder) - other | ARDS | ARDS |
| Sepsis, gynecologic | ARDS | ARDS |
| Sepsis, other | ARDS | ARDS |
| Sepsis, pulmonary - other | ARDS | ARDS |
| Sepsis, unknown | ARDS | ARDS |
| Shock, cardiogenic | nonARDS | nonARDS |
| Shunt-portosystemic, surgery for | ARDS | nonARDS |
| Shunts and revisions | ARDS | nonARDS |
| Skin surgery, other | ARDS | nonARDS |
| Smoke inhalation | ARDS | nonARDS |
| Spinal cord only trauma | ARDS | nonARDS |
| Spinal cord only trauma, surgery for | ARDS | nonARDS |
| Spinal cord sugery, other | ARDS | nonARDS |
| Spinal/extremity trauma | ARDS | nonARDS |
| Spinal/extremity trauma, surgery for | ARDS | nonARDS |
| Spinal/face trauma | ARDS | nonARDS |
| Spinal/face trauma, surgery for | ARDS | nonARDS |
| Spinal/multiple trauma | ARDS | ARDS |
| Spinal/multiple trauma, surgery for | ARDS | ARDS |
| Splenectomy | ARDS | nonARDS |
| Subarachnoid hemorrhage/arteriovenous malformation | ARDS | nonARDS |
| Subarachnoid hemorrhage/intracranial aneurysm | ARDS | nonARDS |
| Subarachnoid hemorrhage/intracranial aneurysm, surgery for | ARDS | nonARDS |
| Tamponade, pericardial | nonARDS | nonARDS |
| Thoracotomy for benign tumor (i.e. mediastinal chest wall mass, thymectomy) | ARDS | nonARDS |
| Thoracotomy for bronchopleural fistula | ARDS | nonARDS |
| Thoracotomy for esophageal cancer | ARDS | nonARDS |
| Thoracotomy for lung cancer | ARDS | nonARDS |
| Thoracotomy for other malignancy in chest | ARDS | nonARDS |
| Thoracotomy for other reasons | ARDS | nonARDS |
| Thoracotomy for pleural disease | ARDS | nonARDS |
| Thoracotomy for thoracic/respiratory infection | ARDS | nonARDS |
| Thrombectomy (with general anesthesia) | ARDS | nonARDS |
| Thrombectomy (without general anesthesia) | ARDS | nonARDS |
| Thrombocytopenia | ARDS | nonARDS |
| Thrombosis, vascular (deep vein) | ARDS | nonARDS |
| Thrombus,arterial | ARDS | nonARDS |
| Thyroid neoplasm | ARDS | nonARDS |
| Thyroidectomy | ARDS | nonARDS |
| Thyroidectomy and parathyroidectomy | ARDS | nonARDS |
| Toxicity, drug (i.e., beta blockers, calcium channel blockers, etc.) | ARDS | nonARDS |
| Tracheostomy | ARDS | nonARDS |
| Transphenoidal surgery | ARDS | nonARDS |
| Transplant, other | ARDS | nonARDS |
| Trauma medical, other | ARDS | ARDS |
| Trauma surgery, other | ARDS | ARDS |
| Tricuspid valve surgery | nonARDS | nonARDS |
| Tumor removal, intracardiac | nonARDS | nonARDS |
| TURP, transurethral prostate resection for benign prostatic hypertrophy | ARDS | nonARDS |
| TURP, transurethral prostate resection for cancer | ARDS | nonARDS |
| Ulcer disease, peptic | ARDS | nonARDS |
| Vascular medical, other | ARDS | nonARDS |
| Vascular surgery, other | ARDS | nonARDS |
| Vasculitis | ARDS | nonARDS |
| Ventricular Septal Defect (VSD) Repair | nonARDS | nonARDS |
| Ventriculostomy | ARDS | nonARDS |
| Weaning from mechanical ventilation (transfer from other unit or hospital only) | ARDS | nonARDS |
| Whipple-surgery for pancreatic cancer | ARDS | nonARDS |

**Table 5**. Scientific Steering Group

| |
|---|
| Dr. Ken Parhar, Principal Investigator MD, MSc[1,2,3] |
| Dr. Andrea Soo, Senior Biostatistician PhD[1] |
| Gwen Knight BA[1] |
| Dr. Sean Bagshaw MD, MSc[5,6] |
| Dr. Kirsten Fiest PhD[1,2,4] |
| Dr. Daniel Niven MD, PhD[1,2,4] |
| Dr. Gordon Rubenfeld MD, MSc[7] |
| Dr. Damon Scales MD, PhD[7] |
| Dr. Tom Stelfox MD, PhD[1,2,4] |
| Dr. Dan Zuege MD, MSc[1,5] |

1) Department of Critical Care Medicine, University of Calgary & Alberta Health Services, ICU Administration - Ground Floor - McCaig Tower, Foothills Medical Center, 3134 Hospital Drive NW, Calgary, Alberta, Canada T2N 5A1

2) Department of Community Health Sciences and the O'Brien Institute for Public Health, University of Calgary, Calgary, Alberta, Canada

3) Libin Cardiovascular Institute, University of Calgary, Calgary, Alberta, Canada

4) Department of Community Health Sciences, University of Calgary, Calgary, Alberta, Canada

5) Critical Care Strategic Clinical Network, Alberta Health Services, Alberta, Canada

6) Department of Critical Care Medicine, Faculty of Medicine and Dentistry, University of Alberta and Alberta Health Services, Edmonton, Canada

7) Sunnybrook Health Sciences Centre, University of Toronto, Toronto Ontario, Canada
